# THE IMPACT OF ROUTINES ON EMOTIONAL AND BEHAVIOURAL DIFFICULTIES IN CHILDREN AND ON PARENTAL ANXIETY DURING COVID-19

**DOI:** 10.1101/2022.03.25.22272950

**Authors:** Vera Lees, Rosie Hay, Helen Bould, Alex S. F. Kwong, Daniel Major-Smith, Daphne Kounali, Rebecca M Pearson

## Abstract

**Aims and hypothesis:** We hypothesised that there would be an association between maintaining a routine during lockdown and both lower emotional and behavioural difficulties in children and lower parental anxiety. We also hypothesised that children of ‘keyworker’ parents would have fewer emotional and behavioural symptoms due to having maintained more normal routines.

**Background:** The Covid-19 pandemic and related public health measures have impacted on mental health of children.

**Methods:** We used data from the Avon Longitudinal Study of Parents and Children (ALSPAC) to explore associations between maintaining a routine, and emotional and behavioural difficulties in children, using linear regression models. We included measures of parental anxiety. We separately explored associations with having a keyworker parent. We used the Carey Infant Temperament Questionnaire and the Revised Rutter Parent Scale for Preschool Children to establish levels of emotional and behavioural difficulties.

**Results:** 289 parents completed questionnaires about their 411 children. Keeping a routine was associated with emotional and behavioural difficulty scores 5.0 points lower (95% CI -10.0 to - 0.1), p=0.045 than not keeping a routine. Parents who reported keeping a routine had anxiety scores 4.3 points lower (95% CI -7.5 to -1.1), p=0.009 than those who did not. Children of keyworkers tended to have lower emotional and behavioural difficulty scores (−3.1 (95%CI -6.26 to 0.08), p=0.056) than children of non-keyworkers. All models were adjusted for relevant potential confounders.

**Conclusion:** Maintaining a routine may be beneficial for both child emotional wellbeing and parental anxiety, although it is also possible that lower parental anxiety levels made maintaining a routine easier. Being the child of a keyworker parent during lockdown may have been protective for child emotional wellbeing.

## INTRODUCTION

There is accumulating global evidence that the ongoing Covid-19 pandemic and associated public health measures including self-isolation, lockdowns and school closures have had a significant negative impact on the mental health of children and young people.^1-3^ There have been increases in mental distress, eating disorders and self-harm amongst children and young people.^3^ Data on younger children is lacking, but our own previous work indicates a rise in emotional and behavioural symptoms in primary school aged and younger children.^4^ It is important to understand factors implicated in such rises, particularly those which could be protective for children and young people. A better understanding of protective factors gives potential to target appropriate intervention to support the mental health of younger children in the event of future pandemics or Covid-19 suppression measures.

One possible protective candidate, which evidence suggests is helpful in children’s daily life, is maintaining routine.^5^ Routine helps us plan, deliver, predict what to expect, and may protect us from anxiety during stressful times.^6,7^ Structure and consistency are important for children; for example, keeping regular mealtimes and bedtimes can help children feel safe and secure.^8^ Predictability and familiarity are especially important for children who are still learning and growing, and has also been shown to be important in the context of adolescent mental health.^9^

Routines have been suggested as an area of focus for intervention among families living in chaotic households with young children who present with behavioural problems and bedtime resistant behaviour.^10^

The Covid-19 pandemic and responses to mitigate its effect had the potential to significantly disrupt routine for both children and their parents. The UK entered its first national lockdown on 23^rd^ March 2020. Lockdown meant that schools were closed to children, other than children of keyworkers and those considered “vulnerable”. The British government specified categories of these “key” or “critical” workers whose work was essential to Covid-19 response, including work in health and social care and other key sectors.^10^ Children defined as “vulnerable” included those on child protection plans with local authorities and those who struggled to engage with education remotely.^11^ For many children during lockdown, their normal routine completely disappeared: their schooling changed, their social contacts, hobbies and childcare changed; and many were no longer able to be in contact with grandparents and extended family. Parents’ routines also changed, with some multitasking home-working with home-schooling or childcare, and others being furloughed from employment. Restrictions were partially lifted with children in nursery, Reception, Year 1 and Year 6 able to return from 1^st^ June 2020. From 15^th^ June 2020, Year 10 and Year 12 were permitted limited contact to help prepare for exams.

One study has found that the pandemic has affected behaviours of children and adolescents, in that it has led to decreased physical activity and disrupted sleep patterns.^12^ The NHS Digital Survey of Mental Health of Children and Young People also demonstrated significant changes for children and young people in terms of their education and usual activities due to the Covid-19 pandemic.^13^

In addition to routines helping children’s mental health, childcare availability and the associated routines may also impact parental mental health, including anxiety.^7^

We investigated whether routine has been protective in child emotional and behavioural difficulties during lockdown, by examining the associations between routine and emotional and behavioural difficulties, and by exploring the impact of Covid-19 on children of “keyworkers”, who may have had a more usual routine, but may have been more worried about their parents being at risk of Covid as frontline workers.

We hypothesised that keeping to routine would be associated with fewer emotional and behavioural difficulties in children and lower anxiety in parents, and that being a keyworker parent would be associated with fewer emotional and behavioural problems in children.

## METHOD

The Avon Longitudinal Study of Parents and Children (ALSPAC) is an ongoing population-based study that recruited pregnant women residing in Avon in the south west of England with expected delivery dates between 1^st^ April 1991 and 31^st^ December 1992.^14-16^ The cohort consists of mothers and their partners (G0) associated with 15,454 pregnancies, resulting in 15,589 foetuses (G1) of which 14,901 were alive at 1 year of age. In 2012, ALSPAC began recruiting and collecting data on the next generation, G2, the children of the G1 participants and grandchildren of the originally recruited G0 women (also known as the ‘children of the children of the 90s’). G2 participants can join the study at any time (from early pregnancy onwards), through an open cohort.^17^ As with the original study, data are being collected from both parents (at least one of whom is a G1 participant) and their children. The study website contains details of all data available through a fully searchable data dictionary (http://www.bristol.ac.uk/alspac/researchers/our-data/).

In June 2020, G1 parents (mean age ∼28 years) completed a questionnaire about each of their G2 children,^18^ and a questionnaire about themselves.^19.^ These questionnaires were completed early in the COVID-19 pandemic (between 26^th^ May and 5^th^ July 2020), as part of ALSPAC’s COVID-19 data collection strategy. Study data were collected and managed using REDCap electronic data capture tools hosted at the University of Bristol.^20^

### Measures of Routine

Participants were asked whether they kept a similar routine (e.g. bedtime, mealtimes) to how things were before the official lockdown was announced on 23^rd^ March 2020, with response options “no, not at all”, “yes, a bit”, “yes, a lot” and, “yes, completely”. The latter two categories were combined into one due to low frequency of participants endorsing the original separate categories, resulting in three categories in total.

### Keyworker Status

G1 parents were asked whether they were a keyworker, or whether their work had been classified as critical to the COVID-19 response, with options “yes”, “no”, or “don’t know”.

### Measures of child emotional and behavioural difficulties

Parents who participated in the COVID-19 survey completed one of two assessments regarding their child’s feelings and behaviour since the lockdown. The assessment version depended on the child’s age. Parents of children less than 36 months old (henceforth referred to as younger children) completed the mood and distractibility subscales of the Carey Infant Temperament Questionnaire (ITQ).^21^ Parents of older children (ages 36 months and older) completed the Revised Rutter Parent Scale for Preschool Children.^22^ Measures which were the same as, or as close as possible to the two used during the pandemic were selected from assessments completed by G1 parents before the pandemic. The same standardised scores were used.^22^

### Measures of parental anxiety

Parental anxiety was measured using the GAD-7, a 7-item questionnaire with scores ranging between 0-21, with scores >10 indicating probable generalised anxiety disorder.^23^

### Covariates

The following covariates were included: maternal age in years (continuous variable); parity (continuous variable); maternal education (binary variable: GCSE equivalent or below, or A level or above); child’s age in years (continuous) and child’s gender (binary).

### Statistical Analysis

Linear regression models were employed, with standardised emotional behavioural problems as the outcome, and routine and keyworker status as categorical exposure variables. Analyses were adjusted for relevant confounding variables for the specific association under investigation considering the relationship with the exposures and outcomes. Relevant variables included child age, gender and maternal education, maternal age, parental anxiety and parity. Amongst the ALSPAC-G2 children, there were 122 sib-sets, and therefore we accounted for clustering using robust standard errors, clustered on parent ID. We also ran linear regression models of the association between maternal anxiety and routine. The relationship between child emotional difficulties and parent anxiety is likely to be complex and bi-directional therefore we did not include the two variables in the same model and rather in this work present the associations with routine. All analyses were adjusted for child age, gender and maternal education, maternal age and parity and were conducted in Stata v.15/MP.

## RESULTS

In total, 411 G2 children had questionnaire data, completed by their 289 parents. Of these 411 G2 children, 214 (52%) were male. The median age for G2 children was 34 months (IQR 16 to 71 months).^17^ Of the 411 children, 404 had complete data on keeping a routine. Of those with data, 41 (10%) did not keep the same routine; 182 (44%) kept the same routine “a bit”, 181 (44%) kept the same routine either “a lot” or “completely”.

Of the 411 children, 328 had complete data including covariates. The analyses reported here concern the children with complete data.

### Routine and Emotional and behavioural difficulties in children

We found a dose-response association between the extent of keeping routine and lower emotional and behavioural difficulties in the children (figure 1). This association was not altered by clustering by maternal ID, or by adjusting for child gender, maternal parity, maternal and child age, maternal educational background (Table 1; higher difficulties scores indicate more emotional and behavioural difficulties). Compared to families where no routine was kept, children of those parents reporting keeping “a lot” or “completely” the same routine to before lockdown had emotional and behavioural difficulty scores 5 points lower (95% CI -10.02 to - 0.12; p=0.045). There was less evidence of a difference in emotional and behavioural difficulties between those reporting keeping the same routine “a bit” in comparison to those not keeping the same routine at all.

**Table 1.** Linear regression comparing the association between the extent of routine and child difficulties score *accounting for clustering according to mother id and adjusted for child gender, number of children, maternal and child age, educational background.

| Routine | Linear regression coefficient for emotional difficulties score (unadjusted) | Linear regression coefficient for emotional difficulties score (adjusted)* |
| --- | --- | --- |
| Not at all | REF | REF |
| A bit the same | -2.13 (95%CI -6.77 to 2.51)<br>$p=0.368$ | -1.59 (95%CI -6.31 to 3.13)<br>$p=0.509$ |
| A lot /Completely the same | -5.62 (95%CI -10.39 to -0.86)<br>$p=0.021$ | -5.01 (95%CI -10.02 to -0.12) $p=0.045$ |

**Figure 1.**
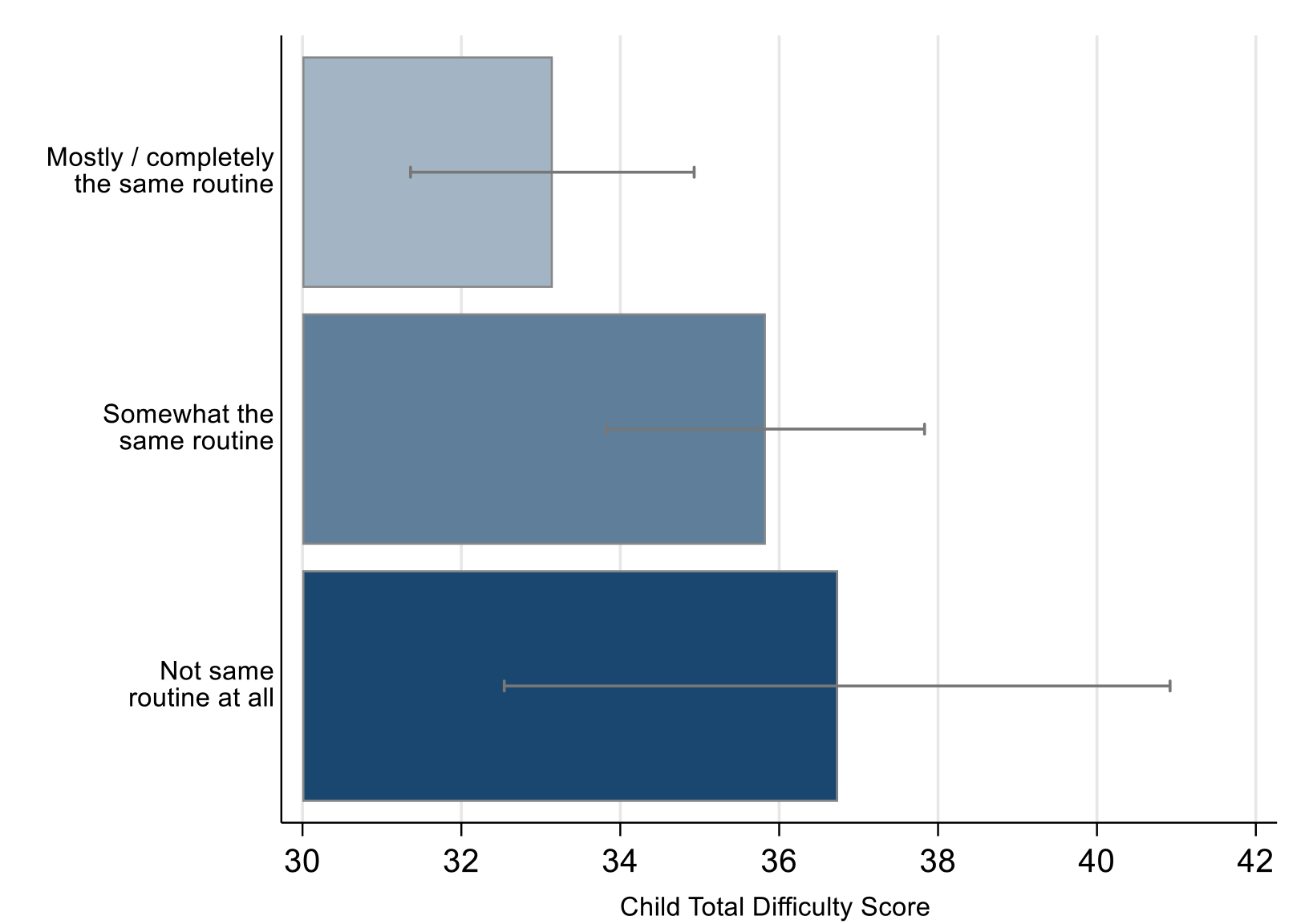
Covid-19 pandemic related total difficulty scores in children by keeping routine in ALSPAC. The error bars represent 95% CI.

### Routine and Parental anxiety

Parents of 314 G2 children had data on anxiety measured in the second COVID-19 questionnaire and on childcare routines. Parents who kept to a routine were less anxious than those who did not, again finding a dose-response relationship (figure 2). A mean GAD score of 11.2 (95% CI 9.82 to 12.51) (indicating probable generalised anxiety disorder) was found in the group that did not follow a routine at all. Parents who kept to the same routine “a bit” had a mean GAD score of 8.9 (95% CI 8.28 to 9.54). Those parents who followed “a lot/completely the same” routine had mean GAD score of 7.5 (95% CI 6.78 to 8.20). Table 2 shows the results of the linear regression analyses describing the association between routine and parental anxiety.

**Table 2.** Linear regression comparing the association between the extent of routine and maternal anxiety scores. *accounting for clustering according to mother id and adjusted for child gender, number of children, maternal and child age, educational background.

| **Routine** | **Linear regression coefficient for maternal anxiety score (unadjusted)** | **Linear regression coefficient for maternal anxiety score (adjusted)\*** |
| --- | --- | --- |
| Not at all | REF | REF |
| A bit the same | -3.46 (95%CI -6.02 to -0.91)<br>$p=0.008$ | -4.43 (95%CI -7.60 to -1.30) $p=0.005$ |
| A lot/Completely the same | -4.30 (95%CI -6.86 to -1.75)<br>$p=0.001$ | -4.30 (95%CI -7.5 to -1.10) $p=0.009$ |

**Figure 2.**
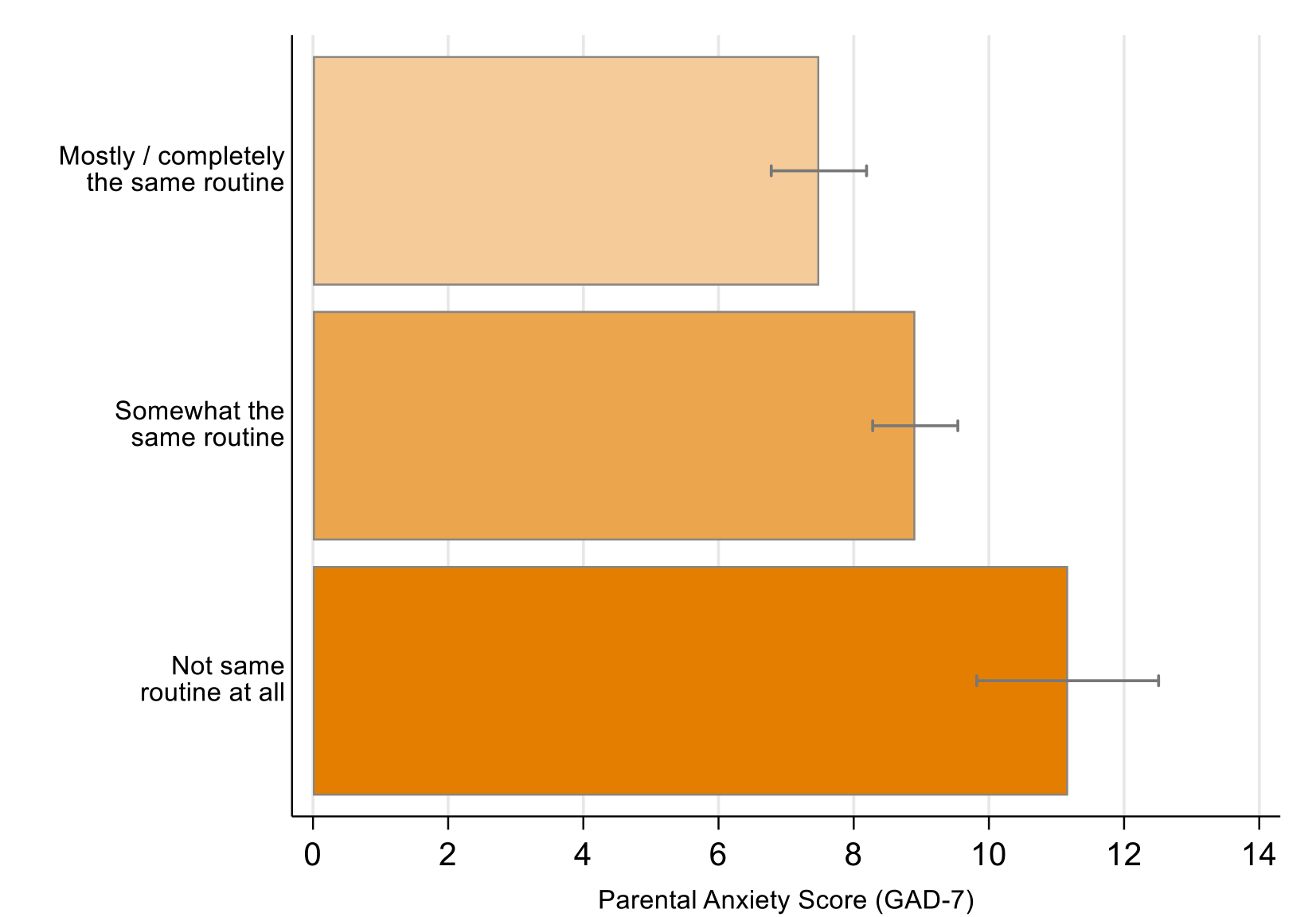
Covid-19 pandemic related parental anxiety score by keeping routine in ALSPAC. Error bars represent 95% CIs.

In the model taking into account maternal and child age, gender, maternal education and parity there was evidence that, compared to families where no routine was kept at all, parents who kept “a lot/completely the same routine” to before lockdown, had anxiety scores on average 4.3 points lower (95% CI -7.5 to -1.1; p=0.009). Those who kept the same routine “a bit” had parental anxiety scores on average 4.4 points lower (95% CI -7.60 to -1.30; p=0.005) than those who did not keep the same routine at all (Table 2).

### Keyworkers

147 children had one or more parents who were keyworkers; and 160 had one or more parents who were not keyworkers. Parents were counted more than once if they had more than one child.

Children with at least one keyworker parent were more likely to have kept similar routine to before lockdown than children with no keyworker parent (Figure 3, (Chi2 (2) 22.13, p <0.001)), whilst they were less likely to report they kept to ‘somewhat same routine’ and no differences were apparent for ‘not at all same routine’.

**Figure 3.**
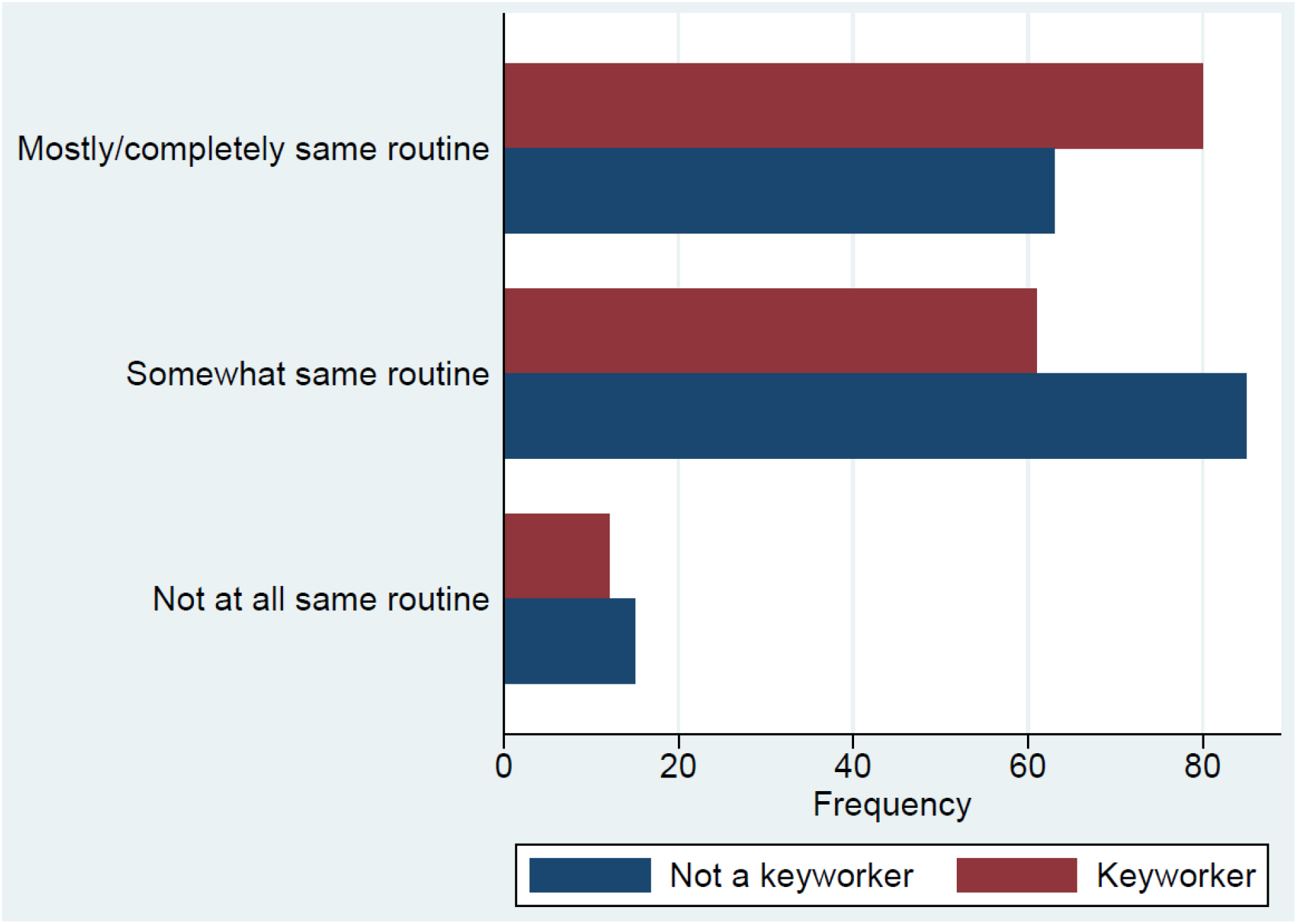
Routine kept by keyworker and non-keyworker parents.

Children of keyworkers were also reported to have fewer emotional and behavioural difficulties during lockdown than children of non-keyworkers (figure 4). After adjusting for maternal and child age, maternal education, gender, and parental anxiety, keyworker children scored on average 3 points lower (95%CI -6.26 to 0.08; p=0.056) than children of non-keyworkers. However, the evidence against an effect of this magnitude occurring under the null hypothesis is weaker, relative to some of our other results. (Table 3).

**Table 3.**
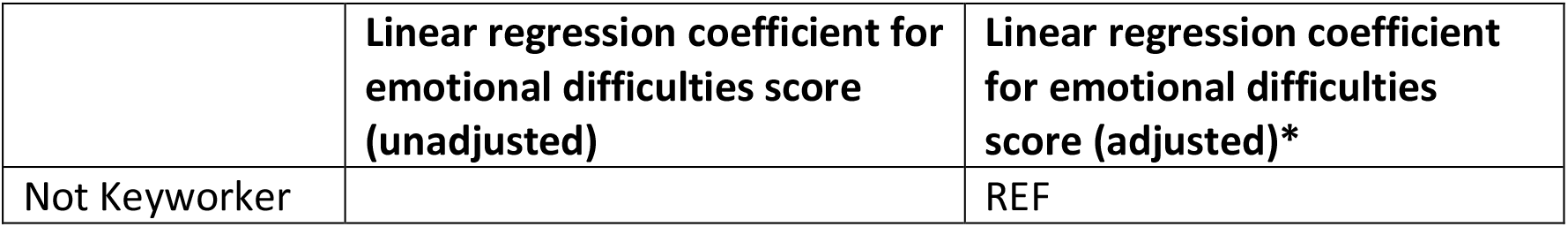
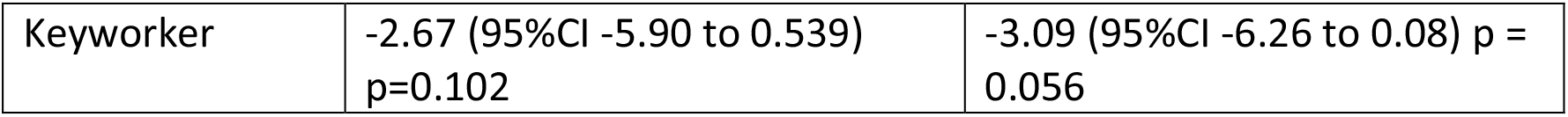
Linear regression comparing the association between parental keyworker status and child difficulties scores. *accounting for clustering according to mother id and adjusted for, number of children, maternal and child age, educational background and anxiety.

**Figure 4.**
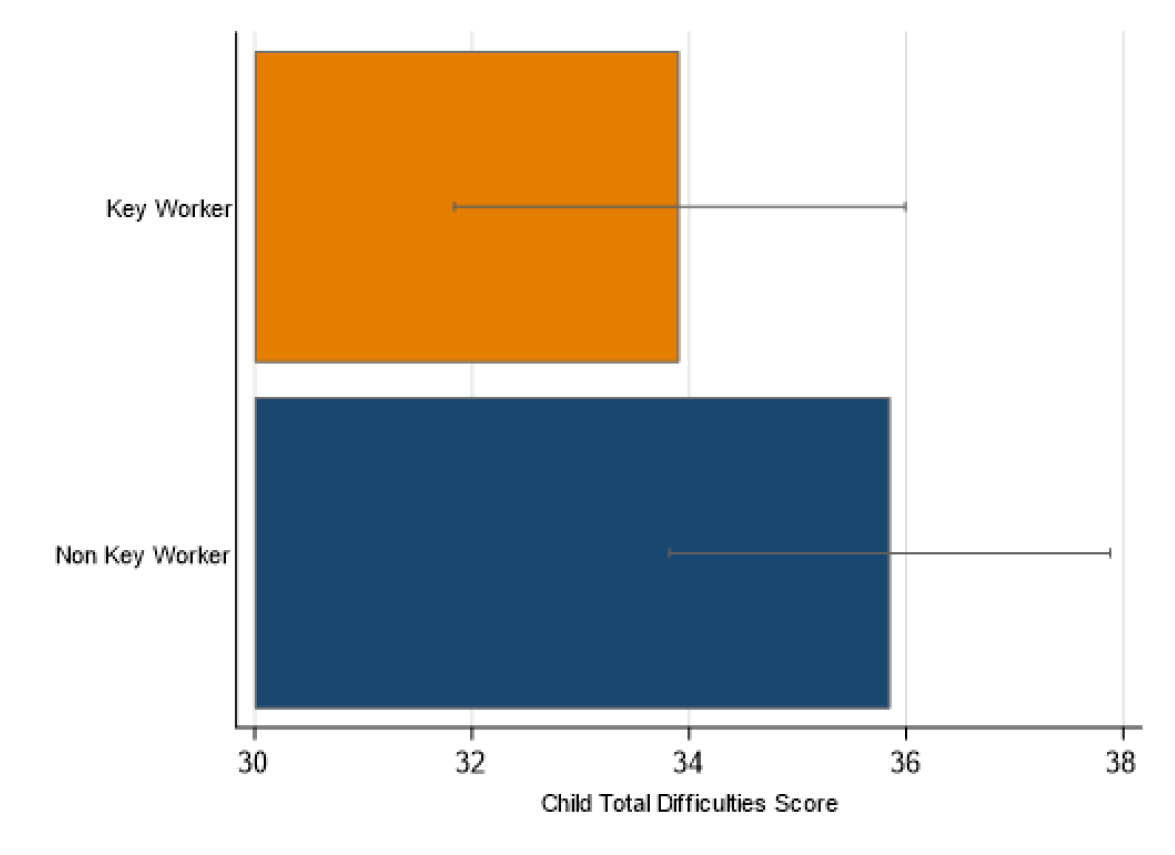
Difficulty scores for children of keyworkers and non-keyworkers. Error bars represent 95% CIs.

## DISCUSSION

We found evidence of an observed dose response between extent to which routine stayed the same and lower emotional and behavioural difficulties scores in children, as well as lower anxiety in parents. We also found that keyworker parents were more likely to keep routines, and a suggestion that emotional and behavioural difficulties scores were lower in children of keyworkers than those of non-keyworkers.

### Strengths and Limitations

This is a novel study looking at the effect of Covid-19 pandemic on younger children, using longitudinal pre- and during-pandemic data from the ALSPAC cohort. Although this provides a good size sample, the population in the sample is limited in ethnic diversity and geographical spread, which may limit the generalisability of our results to other populations. The cross-sectional nature of this work means that, although we have described an association between keeping routine and fewer behavioural and emotional difficulties in children, we are unable to determine the direction of causality, i.e. we cannot conclude that keeping to routine leads to lower emotional and behavioural difficulties, since it is also possible that children having fewer emotional and behavioural difficulties makes it easier to follow a routine. Other possible explanations for these results are that like all cohort studies, ALSPAC has incomplete recruitment and loss to follow-up; the current recruitment at a relatively young age and the focus of analyses in cross-generational effects are more specific to ALSPAG-G2.^16^ Furthermore, whilst we have attempted to adjust for factors that may confound the relationship between routine and emotional and behavioural difficulties, it is possible that residual confounding from unmeasured variables may affect this apparent relationship.

The association between routine and lower parental anxiety is cross-sectional, so we do not know whether routine helps reduce anxiety, or having lower levels of anxiety makes it easier for parents to keep to a routine.

Furthermore, the phrasing of the questions on “routines” compares current routine to pre-lockdown routine. If there was little routine pre-lockdown, then answering “yes, completely” to maintaining routine during lockdown could still indicate lack of routine.

It is not possible to draw conclusions on what specific aspect of routine may be significant in terms of being protective for children from this research. In this paper, routine refers to bedtimes and mealtimes, not wider routines like childcare settings, schooling, exercise/physical activity, and socialising.

Importantly, child emotional and behavioural difficulties are reported by the parent. This is potentially subject to bias. For example, where a parent is struggling with their own mental health, a child’s usual behaviour may be perceived as more challenging because the parent is finding their child’s behaviour more difficult to manage rather than because the child’s behaviour has changed significantly.^25^

### Comparison with other research

As the recent Ofsted report emphasizes,^3^ school closures have resulted in children regressing in their learning as well as increasing mental health distress, eating disorders and self-harm.^3^ Our research further suggests an increase in emotional and behavioural difficulties, associated with difficulties maintaining childcare routines during the same period.

### Implications

We demonstrate that routines are associated with good mental health, both in children and in parents. Whilst we cannot confirm the direction of causality from these data alone, if causal, this would suggest that interventions to facilitate routines could help prevent difficulties. Most parenting advice regarding lockdown mentions keeping routine and this research supports that advice.

Since parents with higher levels of anxiety are less likely to have kept to the same routine, it may be that families need different levels of support to achieve this aim. It is important to support parents’ mental health. Keeping childcare provision open may also mean that routines can be more consistent and may reduce anxiety for parents who are juggling work and childcare responsibilities.

### Conclusion

We conclude that maintaining routine may be beneficial for children’s emotional wellbeing as well as parental anxiety. Further research exploring the direction of causation may provide us with greater understanding. Being the child of a keyworker parent during lockdown may have been protective for child emotional wellbeing. This may be reassuring to keyworkers, however further work to replicate this finding is needed.

## Data Availability

All data produced in the present study are available upon reasonable request to the authors.

## Acknowledgements

We are extremely grateful to all the families who took part in this study, the midwives for their help in recruiting them, and the whole ALSPAC team, which includes interviewers, computer and laboratory technicians, clerical workers, research scientists, volunteers, managers, receptionists and nurses, particularly during this challenging, uncertain and unprecedented period at the beginning of global pandemic.

## Ethics

Ethical approval for the study was obtained from the ALSPAC Ethics and Law Committee and the Local Research Ethics Committees. Informed consent for the use of data collected via questionnaires and clinics was obtained from participants following the recommendations of the ALSPAC Ethics and Law Committee at the time.

